# “A Market-Based Virus Monitoring Strategy Complementary to Epidemiological Surveillance in Urban Settings”

**DOI:** 10.64898/2025.12.22.25342882

**Authors:** Paola Martínez-Duque, Marco A. Jiménez-Rico, Lucio A. Bacab-Cab, Aarón G. Cantón, Rhys P.D. Inward, Bernardo Gutierrez, Sumali Bajaj, Stien Vandendiessche, Azael Che-Mendoza, Olivia Contaldo, James Earnest, Pablo Manrique-Saide, Gonzalo Vazquez-Prokopec, Daniel Canul Canul, Karina Jacqueline Ciau Carrillo, Guadalupe Ayora-Talavera, David Roiz, Henry Puerta-Guardo, Carlos Machaín-Williams, Arturo Reyes-Sandoval, Moritz U.G. Kraemer, Miguel A. García-Knight, Gerardo Suzán, Marina Escalera-Zamudio

## Abstract

Mexico has experienced several viral outbreaks of substantial intensity, including hyperendemic dengue since the mid-1990s, the COVID-19 epidemic in 2020-2022, and recent reports of avian influenza A (H5N1) in poultry, representing an ongoing risk of zoonotic transmission. Under a One Health framework, traditional markets represent valuable sites for virus monitoring at the human-animal interface. As a case study, we used the central market of Merida City in Yucatan, Mexico, a regional hotspot for viral outbreaks. We conducted a pilot evaluation of a virus monitoring strategy drawing from entomological, wastewater, and live bird surveillance. Systematic longitudinal sampling was carried out over 11 months at 1 to 6-week intervals from April 2022 to February 2023. Samples were screened using RT-qPCR, with positives further characterised through sequencing and phylogenetic analyses. Results were interpreted in the context of official national epidemiological data and surveillance efforts. We report an early detection of DENV-3 lineage III-B3.2 (genotype III American II) from a mosquito pool preceding its identification from contemporaneous human-derived sequences derived from genomic surveillance efforts. SARS-CoV-2 RNA was detected from wastewater surveillance throughout both epidemic waves recorded at the city and state level, with a broad overlap across temporal epidemiological trends. Cloacal swabs from two juvenile birds yielded putative signals consistent with detection of avian influenza A virus, suggesting a possible cryptic virus circulation in live birds sold at the market. Our study demonstrates the feasibility of implementing similar strategies to monitor multiple viral pathogens in urban settings. While not suitable for the detection of novel outbreaks, this approach may complement existing surveillance efforts by delivering molecular signals supporting the detection of epidemiologically relevant viruses in circulation, particularly in settings where genomic capacity remains limited.

## INTRODUCTION

Viral pathogens are a major threat to global health security. During the COVID-19 pandemic, the SARS-CoV-2 virus alone resulted in at least seven million deaths worldwide [1] and an estimated economic loss exceeding 10% of the global GDP [2]. Other viruses causing recurrent epidemics, including dengue (DENV) and influenza A, are a persistent burden on economies and global health [3,4]. In this light, effective virus surveillance strategies are essential, but must account for disparities in resource availability and sequencing capacity across regions [5–8].

Traditional markets are retail places where live animals and their products are sold, representing ecological niches where humans converge with populations of reservoir animals, vectors, and the viruses that they carry [9]. Moreover, the animal-human interface intrinsic to markets may increase the risk of pathogen transmission (extensively reviewed in [10]). As an example, recurrent H5N1 outbreaks in poultry sold at markets show that repeated human to animal contact facilitate spillover events [11,12], whilst trade related to live-animal markets is a key predictor of global zoonotic pathogen richness [13]. Moreover, some studies have identified these sites as potentially increasing the risk of transmission for mosquito-borne diseases in tropical and subtropical regions, as places where mosquito breeding conditions and human activity coincide [14]. In this context, traditional markets offer an opportunity for virus monitoring, particularly in regions where genomic surveillance could be strengthened by additional molecular data [10,15–17].

Mexico represents a geographic, cultural, and economic corridor between Central America and the United States, possibly associated with virus spread across regions [18–21]. The country has historically faced multiple viral epidemics, consistently reporting some of the highest incidence rates of dengue in Latin America [22], and with seasonal influenza A infections frequently reaching epidemic levels [23,24]. Mexico was also one of the most severely impacted countries by SARS-CoV-2 in the Americas [21], and was the location where the 2009 influenza A pandemic virus (pdmH1N1) was first identified [18]. Moreover, recent reports of avian influenza A virus (AIV) poultry infections across the country [25] underscore the need to strengthen surveillance strategies developed under a One Health framework [5,6], [26,27].

In the country, infectious disease surveillance is coordinated by the National Institute for Epidemiological Diagnosis and Reference (InDRE) and the National Epidemiological Surveillance System (Sistema Nacional de Vigilancia Epidemiológica, SINAVE) [28], integrating case reporting from healthcare facilities nationwide [29]. With hyperendemic dengue (*i.e.,* displaying a continuous and widespread circulation of all four serotypes) representing a major epidemiological concern across broad geographic regions [22], national surveillance efforts must rely on a sentinel approach [28]. This includes confirmed diagnosis and virus serotyping of around 25% of all reported symptomatic cases, centralised at the National Arbovirus Reference Laboratory-InDRE, a WHO-accredited reference laboratory [30].

Additional surveillance components include vector population monitoring implemented nationally since 2013 [31], integrating climatic, entomological and epidemiological data into an early warning alert system (EWARS) [32,33]. Recent advances in retrospective genomic surveillance achieved in collaboration with national public health institutions also show promise in strengthening public health decision-making [19]. However, the gap between reported cases and available viral genomic data remains [22,34] (for an extended contextualisation of recent DENV epidemiology and dynamics in Mexico, see **S1 Text** and **S1 Fig)**. Together with a limited molecular characterisation of circulating viruses, this may include overlooking the introduction of virus lineages circulating in neighbouring regions that can lead to new outbreaks [35,36].

The state of Yucatan, located in south Mexico, is hub for the introduction and transmission of several viruses, including SARS-CoV-2, DENV, chikungunya (CHIKV), and Zika (ZIKV) [19,20,37,38]. Here, we present a proof-of-concept pilot study assessing whether a targeted virus monitoring approach implemented at a traditional market can capture molecular signals of viral circulation across urban settings. Using as case study the Central Market of Merida City (the state’s capital), we implemented a monitoring strategy to detect multiple viruses labelled as of ‘epidemiological priority’ by the Ministry of Health Mexico: SARS-CoV-2, Influenza A and mosquito-borne flaviviruses (*i.e.,* DENV, CHIKV and ZIKV). Samples derived from entomological, wastewater, and live bird monitoring were characterised through sequencing and phylogenetic analyses, with results interpreted in the context of official epidemiological data [28].

We report the identification of DENV-3 III-B (genotype III American II lineage) in mosquitoes prior to its detection in available contemporary genomic data from human cases from the region [39]. Monitoring SARS-CoV-2 RNA from wastewater resulted in positive signals aligning with the two infection waves officially reported both at a city and state level. Finally, we identify two putative positive samples to avian influenza A (AIV), suggesting a possible cryptic virus circulation in the bird population sold at market. Our findings suggest that even under sparse sampling, the detection of viruses of epidemiological relevance at this site was feasible. The added value of the approach presented lies in generating additional molecular signals of virus circulation directly from environmental and animal samples. In this light, localised virus monitoring using molecular approaches may complement routine surveillance efforts.

## METHODS

### Study Area

The study area comprised the San Benito (20.9681°N, –89.6299°W) and Lucas de Galvez (20.9636°N, –89.6207°W) public markets, located within the historic centre of Merida City, Yucatan, Mexico. These facilities form a single interconnected market area, sharing a common parking site with an on-site local wastewater treatment plant. Together, they represent the largest commercial hub in the city, including butcheries, produce vendors, and animal vending stalls, and well as multiple small restaurants. According to data from the TIRS trial (Targeted Indoor Residual Spraying; a large cluster-randomised trial conducted in Merida city during 2022-2023) [40], the market area is also located within a hot-spot area for dengue cases in Merida City.

### Mosquito surveillance

Mosquito sampling was conducted in the shared basement parking level, where the wastewater treatment plant is located. Conditions create a stable microenvironment for mosquito breeding and resting. Thus, the area provided a consistent sampling environment with higher expected mosquito density. Throughout a total of 13 sampling periods, five Biogents BG-Sentinel traps (Biogents, Germany) baited with BG-Lure attractants were deployed. Traps were spaced 30-50 m apart to avoid competition, generally operating in 72-h cycles, equivalent to 360 trap-hours per cycle. Two exceptions occurred in April (2 days, 240 trap-hours) and May (6 days, 840 trap-hours) while protocols were established. In total, sampling effort comprised 4,320 trap-hours (180 trap-nights, defined as one trap operating for 24 h) (**S1 Table**) [41].

At the end of each 24-h cycle, collection bags were replaced, transported to the laboratory at 4°C, and stored at –80°C until further processing. Mosquitoes were taxonomically identified using a stereomicroscope (Leica Microsystems, Singapore) and the Clark-Gil and Darsie morphologic key for mosquitoes in Central America [42]. Specimens were placed into 1.5 mL microtubes, with initial pools constructed by species, sex, feeding status, collection date, and trap number (1-50 individuals per pool). *Aedes aegypti* was prioritised for subsequent virological analysis given its role as a primary vector of epidemiologically relevant viruses endemic circulating within the region (*i.e.,* DENV and CHIKV). The *Culex quinquefasciatus* population, comprising the majority of individuals collected, was down-sampled to ensure a feasible screening effort while retaining species representation. From the total Culex females collected, approximately 30% were selected and grouped into 164 batches, including 65 blood-fed/gravid females processed individually and 99 pooled samples. All *Aedes* females were processed in 222 batches, including 86 blood-fed/gravid females processed individually and 136 pooled samples (**S1 Table**). A total of 386 batches were homogenised using sterile pestles in Dulbecco’s Modified Eagle Medium (DMEM), with volume adjusted according to pool size. Homogenates were centrifuged at 14,000 rpm for 3 min, with 140 µL of supernatant retained for RNA extraction and stored at –80°C.

### Wastewater surveillance

Wastewater sampling was conducted at the basement parking area of San Benito, where a self-contained water treatment plant that releases and recycles non-potable treated water for both market areas. The plant consists of two units: U1 receives sanitary waste, while U2 receives food waste via a grease trap. Pre-treatment flocculation tanks from each unit served as independent collection points for daily sampling (Monday to Friday; 50 mL per point). Samples were transported at 4°C, centrifuged at 4,200 rpm (3,500 x *g*) for 30 min at 4°C, and separated into 500 μL sediment and 1.2 mL sludge/supernatant. Sludge aliquots were mixed 1:1 with RNA Shield (Zymo Research, USA). Composite samples were generated by combining equal-volume daily aliquots within each epidemiological week and homogenised prior to extraction. Samples comprising a single 140 μL fraction were used as input for RNA extraction, with no additional filtration performed. All samples were stored at –80°C until processing, following recommended guidelines for wastewater surveillance [43].

### Bird monitoring

Live bird sampling at the animal vending facilities at Lucas de Galvez was conducted once (May 17^th^ 2022) during the initial pilot phase of our program. Sampling represented 30% (n= 21/70) of all birds within accessible cages (**S3 Fig**), in line with exploratory strategies following a low-to-moderate virus prevalence scenario [44]. With consent and authorization of stallholders, cloacal swabs were collected from individuals representing different avian species: juvenile turkeys (*Meleagris gallopavo,* n=5), chicks (*Gallus gallus domesticus,* n=9), juvenile and adult ducks (*Anas platyrhynchos domesticus,* n=5), guinea fowl (*Numida meleagris,* n=1), and one goose (*Anser anser domesticus,* n=1). Swabs were placed in cryotubes containing DNA/RNA Shield (Zymo Research, Irvine, CA, USA), transported at 4°C, and stored at –80°C until further processing. Additional sampling could not be performed due to administrative restrictions imposed by the market’s authorities. An interactive visualization of the animal vending facility layout with photographs of sampled cages is available in **S1 File**.

### RNA extraction and cDNA synthesis

All samples were thawed at 4°C and processed for RNA extraction using the QIAamp Viral RNA Mini Kit (QIAGEN, Germany) using linear acrylamide as a carrier and eluting in 60μL of PCR-grade water. Extracted RNA was used directly for all RT-PCR-based detection assays. In parallel, complementary DNA (cDNA) was synthesised using the SuperScript IV First-Strand Synthesis System (Invitrogen, USA) with random hexamers. Purified cDNA was eluted in 20μL of PCR-grade water and stored at −20°C as a stable archive to account for potential RNA degradation.

### Arbovirus screening

Pan-flavivirus PCR screening was done using two nested RT-PCR assays targeting non-overlapping conserved regions of the NS5 gene [45,46]. These protocols can detect a broad range of flaviviruses, including Aedes-associated viruses (DENV and ZIKV), as well as Culex-associated viruses (West Nile Virus [WNV] and St. Louis Encephalitis Virus [SLEV]). Screening was applied to both mosquito as well as avian samples. The protocol described by Sánchez-Seco et al. [46] was initial used, targeting a short 143 nt fragment. The protocol described by Vázquez et al. [45] was then used to confirm positive samples, targeting a longer 1,019 nt fragment. Both protocols were implemented without modification: the first PCR round was performed using the QIAGEN OneStep RT-PCR Kit (QIAGEN, Germany), whilst the second PCR was performed using the HotStarTaq Plus DNA Polymerase (QIAGEN, Germany). Each 25μL reaction contained 2.5μL of RNA template, which was amplified for 40 cycles using the SimpliAmp Thermal Cycler (Applied Biosystems, Thermo Fisher Scientific, USA). Nuclease-free water was used as a negative control, and a clinically-derived sample confirmed for dengue was used as a positive control.

Following initial screening, positive samples were further tested for specific viruses (DENV, CHIKV, and ZIKV). This targeted approach was applied only to mosquito pools, as positive detections only originated from these samples. PCR screening was done using the Genesig® RT-qPCR multiplex kit (Primerdesign Ltd., UK), targeting a conserved genomic region of each virus, designed to detect >95% of sequences available in the NCBI database up to March 2023 [47]. Each 20 μL reaction contained 5μL of RNA template and was amplified for 50 cycles using the CFX96 thermal cycler (Bio-Rad, USA). Positive and negative controls supplied with the kit were included in each run. A Ct value <40 was considered positive, following the manufacturer’s specifications.

### Respiratory virus screening

Wastewater samples were screened using the Allplex Assay RT-qPCR kit for detecting SARS-CoV-2, FluA, FluB, and the RSV viruses (Seegene, South Korea). In addition to targeting highly conserved genomic regions of these respiratory viruses, the assay simultaneously amplifies three SARS-CoV-2 gene regions (S, RdRP, and N), incorporating both endogenous and exogenous internal controls to ensure assay validity. Each 20μL reaction contained 5μL of RNA template amplified for 45 cycles using the CFX96 thermal cycler (Bio-Rad, USA). Positive and negative controls provided with the kit were included in each run. For SARS-CoV-2, an additional positive control (2019-nCoV N PC, IDTdna) was included. A Ct value <40 was considered positive, as specified in the kit’s documentation.

### Influenza A virus screening

Screening for influenza A viruses in cloacal swabs from live birds was done using the Genesig® Influenza type A M1 qPCR kit (Primerdesign Ltd., UK), which amplifies a 244 nt fragment of the highly conserved M virus gene segment [48]. The kit has been successfully employed to detect both human (IAVs) and avian influenza A virus (AIVs) in multiple sample types [49]. Positive samples were then subtyped using the Genesig® Influenza H5 and H7 Kits (Primerdesign Ltd., UK). Each 20μL reaction contained 5μL of RNA template amplified for 50 cycles using the CFX96 thermal cycler (Bio-Rad, USA). Positive and negative controls provided in the kits were included in each run. A Ct value <40 was considered positive, as specified in the kit’s documentation.

### Sequencing

PCR amplicons were visualised on 2% agarose gels stained with SYBR Safe (Thermo Fisher Scientific, USA). Selected PCR products according to expected band sizes were purified using the ExoSAP-IT Express PCR Product Cleanup Kit (Thermo Fisher Scientific, USA) and sent to Macrogen Europe (Netherlands) for direct sequencing. Sequencing was carried out using the BigDye Terminator v3.1 Cycle Sequencing Kit (Applied Biosystems, USA) on an ABI 3730xl DNA Analyzer. Low-quality base calling sequence ends were trimmed, primer sequences were removed, and forward and reverse reads were assembled to generate an overlapping consensus sequence. Outputs were processed to generate consensus sequences (FASTA format) using Geneious Prime v.2023.1.2 (Biomatters Ltd., New Zealand). Sequence homology was assessed using BLAST, employing both blastn (for more divergent sequences) and megablast (optimised for highly similar sequences) to search against the NCBI GenBank database (as of March 2023) [47]. All hits returned were inspected to avoid overlooking more divergent but potentially relevant matches. A cut-off threshold of ≥90% identity and an E-value 1×10E-5 was then used as a post hoc conservative criterion to prioritise high-confidence matches [50]. For more divergent sequences, we also explored alternative BLAST parameter, including a relaxed reward/penalty ratio of about one (1/-1) best for sequences that less conserved (<75% pairwise identity) [51].

### Virus isolation and serotyping

Following RT-qPCR identification of DENV-3 in a mosquito pool, RNA directly extracted from the original mosquito homogenate was used as template for PCR. To increase detection sensitivity, virus isolation was also performed from RNA extracted from infected cell culture. For this, the original sample was clarified by centrifugation at 10,000 *g* for 10 min at 4°C. The supernatant was filtered through a 0.22 μm membrane before inoculation on a confluent monolayer of Vero cells (DMEM + 2% FBS, 37°C, 5% CO₂) and C6/36 cells (DMEM + 10% FBS, 28°C, non-CO_2_ incubator). Cell culture was monitored for cytopathic effects (CPE), with a second passage performed to confirm infection. DENV RT-qPCR of cell culture supernatants at 2 days post-infection (dpi) showed decreased Ct values (from 30.93 in the original homogenate to 29.38).

Amplification of a larger E gene fragment from our mosquito pool was unsuccessful within a single PCR round. Thus, this required us to develop a semi-nested PCR approach to generate sufficient material for sequencing and subsequent analyses. Based on a DENV-3-specific set of primers used for whole viral genome amplification [19], our semi-nested PCR assay was developed to amplify a fragment of the envelope (E) viral gene. The first PCR round (R1) amplifies a 951 nt fragment (P02f: 5′-GGGAGTAGGAAACAGAGATTTTGTGG-3′, and P02r: 5′-GAGTATTGTCCCATGCTGCGTT-3′. The second PCR round (R2) amplifies a 589nt fragment (P03f 5′-ACCAATAGAGGGAAAAGTGGTGC-3′, P02r). Both reactions were carried out using with the NEB Q5 High-Fidelity 2X Master Mix (New England Biolabs, USA) with the MultiGene OptiMax thermal cycler (Labnet International Inc., USA). Cycle conditions for R1 were as follows: 98°C X 30s; 35 cycles at 98°C X 10s, 68°C X 30s, 72°C for 1 min and final extension at 72°C X 2 min. For R2: 98°C X 30s; 35 cycles at 98°C X 10s, 68°C X 20s, 72°C X 30s and final extension at 72°C X 1 min. Nuclease-free water was used as a negative control, and a clinical RNA sample previously confirmed as dengue-positive human case was used as a positive control.

### Phylogenetic analysis

We compiled a dataset comprising 1243 high-quality DENV-3 genomes (>80% coverage) corresponding to all human isolates publicly available on the NCBI GenBank database as of August 2025 [47]. The dataset represents full DENV-3 Clade III diversity circulating in North, Central and South America sampled between 2000 to 2024, including eight publicly available genomes from Mexico from 2003 to 2021 [19], and six additional genomes from Yucatan sampled in 2023 (data source: *Puerta-Guardo et al. unpublished*. Average genome coverage: >90%, Average depth: 20X) (**S2 File**). The later correspond to DENV-3 sequences generated from human cases identified during the TIRS trial (Targeted Indoor Residual Spraying, a large cluster-randomised trial conducted in Merida city during 2022-2023) [40]). This data was shared by collaborators specifically for the purposes of this study, but has not yet been made public.

A Multiple Sequence Alignment (MSA) was generated using MAFFT v7.5 [52] (*mafft – auto*), excluding our partial DENV-3 sequence. A maximum likelihood (ML) phylogenetic tree was then reconstructed using IQ-TREE multicore version 2.2.6 (command*: –nt 8 –m GTR+I+G –B 1000 –keep-ident*) [53]. The overall topology (*i.e.,* clade positioning and ancestry descendance relationships) of our ML tree was validated by comparing it to the global Nextstrain DENV-3 phylogeny [54]. The reference tree was rooted using three selected DENV-3 genomes assigned to the DENV clade 3II-A collected in 2007-2008 (FJ639716, GU131939, and GU131912) identified as basal to DENV-3 clade 3III within the Nextstrain phylogeny. Phylogenetic placement of our partial DENV-3 sequence within the reference tree was performed using EPA-ng (Evolutionary Placement Algorithm) [55] with outputs analysed using GAPPA (Genome And Phylogeny Placement Analysis) [56]. EPA-ng implements a ML framework to place query sequences onto a fixed topology, reducing potential artefactual placements of short sequences [55]. In parallel, we performed full phylogenetic reconstruction by incorporating our DENV-3 partial sequence within the full MSA alignment (command*: –addfragments)* in MAFFT v7.5 [52]. We then re-estimated the phylogenetic tree using IQ-TREE [53] (command*: –g*) constraining the search algorithm to the reference topology. The resulting tree was time-scaled using TreeTime [57], re-estimating branch lengths based on tip sampling dates. We used a fixed molecular clock rate of 1×10^-3^ substitutions/site/year, consistent with previous estimates for global DENV evolution [58]. An interactive visualization of the ML tree with time-stamped, geolocated data can be accessed in **S3 File** (https://microreact.org/project/wSZWLKdUjeHfnDgaBwkU1n-denv-3).

### Ecological characterization of mosquitoes

Mosquito population structure was characterised by estimating absolute and relative abundances and sex ratios for the species collected over the sampling period. Absolute abundance is defined as the number of individuals per species captured across each sampling period. Relative abundance corresponds to the proportion of individuals from each species in the total population sampled [59]. Sex ratio is expressed as the number of males to females within a given population [41]. Infection rate was estimated using both the Minimum Infection Rate (MIR) and Maximum Likelihood Estimation (MLE), two standard approaches for analysing pooled vector samples [60,61]. MIR was defined as the number of positive pools per 1,000 mosquitoes tested, assuming a single infected individual per positive pool. As MIR may underestimate infection rates [60], MLE was additionally used to estimate infection rate obtaining 95% confidence intervals under a binomial likelihood [61]. Both MIR and MLE were computed using the function *pooledBin()* in the binGroup R package (v4.3.2) [62–64].

### Climatic data and GLM analysis

In line with the EWARS framework applied in Mexico for entomological monitoring [33,65], climatic data was obtained from the Merida Atmospheric Observatory, part of the University Network of Atmospheric Observatories (RUOA) of the National Autonomous University of Mexico (UNAM) [66]. Variables included geometric mean temperature (GMT, °C), mean minimum temperature (Tmin, °C), mean maximum temperature (Tmax, °C), mean diurnal temperature range (DTR, °C), cumulative relative humidity (RH, %), and cumulative (aggregate over the time periods specified) precipitation (Ppt, mm) (**S3 File**). The geometric mean was used for temperature. As biological rates respond multiplicatively to temperature fluctuations, this metric a better summary of thermal conditions compared to the arithmetic mean. Other climatic variables (Tmin, Tmax, DTR, RH, precipitation) were summarised using arithmetic means or cumulative values because their expected additive effects.

To explore environmental predictors of local mosquito population dynamics, we used Generalised Linear Models (GLMs) to evaluate associations between mosquito abundance and climatic variables. We modelled mosquito abundance under a negative binomial distribution, stratifying populations by species and sex across four temporal windows (7, 14, 21, and 28 days) [67]. Prior to model fitting, initial data exploration was conducted to assess collinearity among predictors and to identify outliers using the Variance Inflation Factor (VIF, thresholds: >5) (**S2 Table**). A conservative threshold was applied to exclude autocorrelation across predictors, following recommendations for ecological modelling using environmental data [68]. Because RH and Ppt showed strong collinearity, separate GLMs were built using either RH or Ppt along a unique temperature variable. All models were fitted using the *glm.nb* function part of the MASS package using R (v4.3.2) [69]. Best-fit model selection was guided by Akaike Information Criterion (AIC) ranking, whilst overall model fit was evaluated using standard goodness-of-fit diagnostics to ensure meaningful explanatory power. The deviance explained/pseudo-R^2^ was used to quantify the contribution of climatic predictors to mosquito abundance. Coefficients were considered significant at p < 0.05. Model outputs are available in **S4 File**.

### Epidemiological data analysis

Case data for dengue and COVID-19 from 2022-2023 from both Merida city and wider Yucatan state were obtained from official records from the Ministry of Health Mexico [30]. Following the official epidemiological calendar, data were aggregated weekly at the municipal and state level. The temporal association between mosquito abundance and new dengue cases reported monthly was assessed using linear regression and Spearman’s rank correlation. Given that trap effort remained relatively consistent across all sampling periods (excluding the April and May records to preserve temporal comparability), absolute abundance was used as a proxy for mosquito density reflecting temporal variation in population size.

To examine the relationship between SARS-CoV-2 RNA detection in wastewater and COVID-19 cases reported, we modelled the probability of wastewater positivity as a function of weekly new case counts in Merida City. Given a limited number of SARS-CoV-2-positive samples, genome copy number could not be employed [70,71]. Instead, the association between new cases and positive samples was evaluated using logistic regression. For this, we modelled the probability of positive samples as a function of weekly case counts, fitting a Firth-penalised logistic regression to account for sparse sampling [72]. Weekly case counts were unscaled. For interpretability, odds ratios were presented per 500-case increase, using the following transformation: OR_500=exp(βx500). All analyses were performed with the *logit* and associated statistical functions in R (v4.3.2) [73].

## RESULTS

We implemented a pilot program to evaluate the potential of Merida City’s central market area as a sampling site for virus monitoring. Longitudinal sampling was carried out across the central market location, encompassing three main components: (i) the basement parking area (San Benito) for entomological monitoring, (ii) the water treatment plant (San Benito) for wastewater monitoring, and (iii) the animal vending facilities (Lucas de Galvez) for live bird monitoring (**Fig. 1a, S3 Fig**). Samples were screened using RT-qPCR for known viruses of epidemiological relevance in Yucatan circulating earlier that year or in previous years [74]. For mosquito and bird samples, we performed a pan-flavivirus screening, followed by a targeted detection of DENV, ZIKV and CHIKV [19,75]. For wastewater samples, this screened for SARS-CoV-2 [20], influenza (human FluA/FluB [76,77]), as well as the respiratory syncytial virus (RSV, included in the kit. No local epidemiological data available and no positive samples detected). For bird samples, additional screening for avian influenza viruses (H5/H7) [76,77] was performed (**Fig. 1b**). The pilot ran from late April 2022 to early February 2023, with a systematic sampling every 1-6 weeks across 11 months and a frequency varying according to sample type (**Fig. 1c**). Although official data on market’s human mobility was not available, moderate to high human activity was observed throughout the study period. Human activity level in the market varied markedly throughout the day, with peak foot traffic in the early morning and around midday, and reduced activity in the late afternoon. The market operates year-round, with seasonal variation expected with increased movement during local holidays.

**Fig 1.**
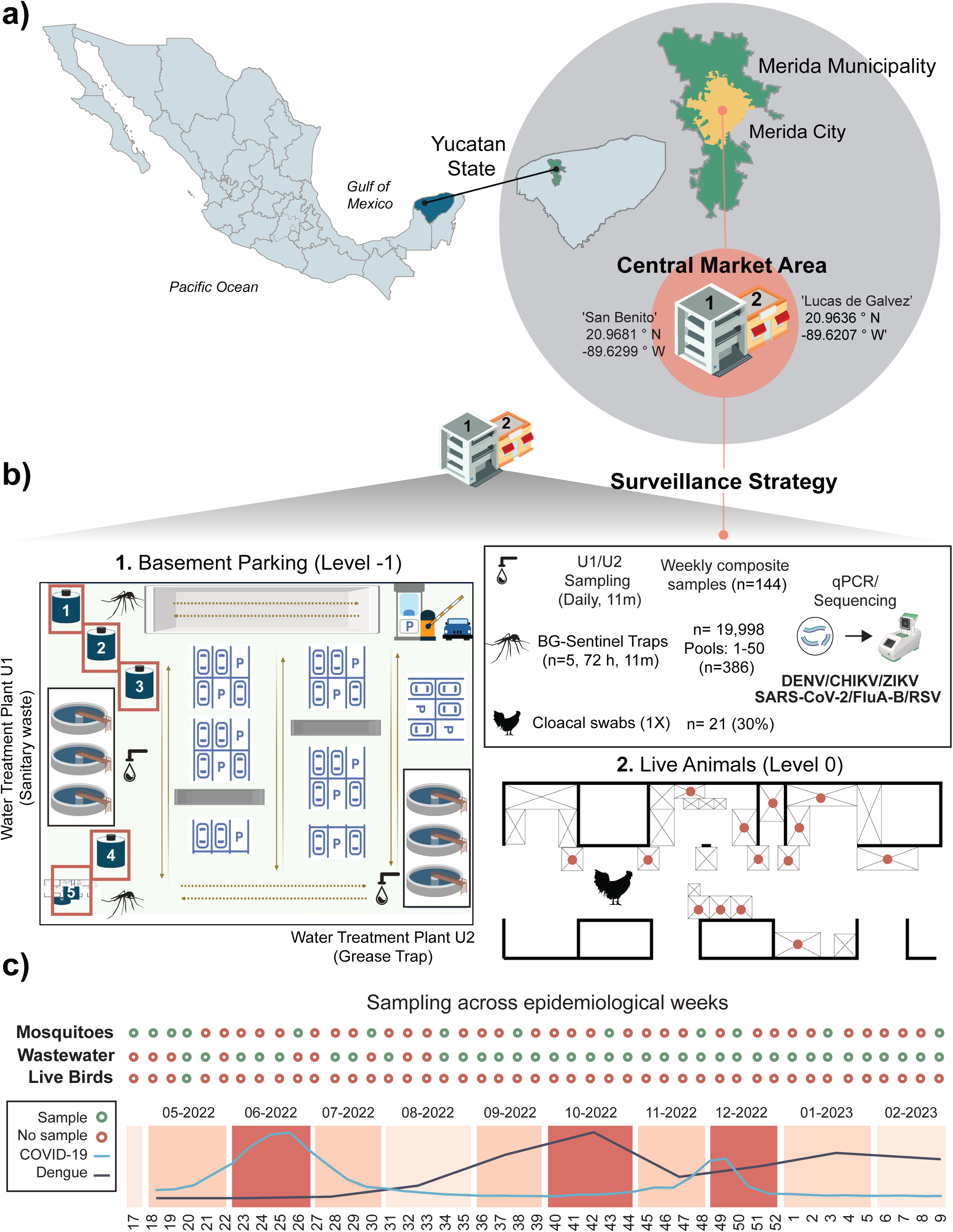
Targeted virus surveillance at the central market of Merida City, Yucatan. (a) Map of Mexico highlighting Yucatan state, showing the location of Merida City and its central market complex as our main sampling site: the Lucas de Galvez and San Benito market areas. (b) Schematic overview of the surveillance strategy comprising three components: (1) entomological surveillance within the basement parking of San Benito, where five BG-Sentinel traps were deployed for 72 hours at 11 m below street level to capture mosquitoes; (1) wastewater surveillance from the market’s treatment plant (U1 = sanitary wastewater; U2 = food waste), where daily samples were taken and pooled weekly; and (2) live bird monitoring at the animal vending facilities (Lucas de Galvez), where cloacal swabs were collected once from approximately 30% of the total of birds within the facilities. (c) Temporal summary of sampling activities across the official national epidemiological calendar. Green circles indicate epidemiological weeks were sampling took place. Red circles indicate weeks without sampling. Sampling periods overlapped with epidemic waves reported COVID-19 and dengue in Yucatan state during 2022-2023. An increase in new case counts is shown by colour gradients (light pink = lowest case count, red = highest case count).

### Entomological surveillance: composition of mosquito populations

The basement parking area was selected for mosquito sampling based on ecological and operational criteria, including persistent shade, high humidity, limited ventilation, and the presence of the local wastewater treatment plant. These conditions create stable breeding and resting conditions for mosquito populations. Moreover, the basement area is structurally connected to both markets and experiences a considerable human activity, making it a likely hotspot for mosquito abundance. Taxonomical characterization of mosquitoes revealed two main species co-occurring in the market: *Culex quinquefasciatus* (72%, n=14,357/19,998) and *Aedes aegypti* (28%, n=5,641/19,998). From all captured *Ae. aegypti mosquitoes*, 62% (n = 3,448) were females, and 37.3% (n = 2,107) were males. From all captured *Cx. quinquefasciatus* identified, 64% (n = 9,086) were females and 36% (n = 5,206) were males. In this context, the sampled mosquito population represents a mix of newly emerged, host-seeking, and blood-fed or gravid individuals (**S1 Table**). Absolute mosquito abundance revealed population fluctuations over time, with both species peaking around September 2022, followed by a gradual decline towards the end of the year. *Cx. quinquefasciatus* displayed two additional peaks in December 2022 and February 2023. Relative mosquito abundance stratified by sex revealed a higher female abundance for *Ae. aegypti* during April 2022, July 2022, October 2022 and February 2023. For *Cx. quinquefasciatus*, a higher female abundance was observed during April 2022, September 2022 and January 2023 (**S4 Fig**). The presence of local breeding sites may result in a proportion of nulliparous mosquitoes, which could reduce the probability of arbovirus detection. Nonetheless, our sampling outputs are consistent with female-biased captures typically reported for BG-Sentinel traps, with both species and sexes sufficiently represented to characterise population composition [78].

### Climatic variations and local mosquito populations

In light of the existing national vector monitoring program and the EWARS framework supporting dengue surveillance in Mexico [30,32,65], we contextualised our entomological findings using local climatic data. For *Ae. aegypti*, we identified precipitation, relative humidity, and temperature at 21-28-day lags as climatic predictors of mosquito abundance within the market (ΔAIC < 2). In general, models incorporating precipitation or relative humidity showed better fit and explained a greater proportion of temporal variation in mosquito abundance than temperature metrics alone. The best-fit models (M208, M220, M196; AIC = 144.6-145.0; deviance explained: 48.1-56.7%) showed positive associations for precipitation (β = 0.019 ± 0.007, p = 0.004), relative humidity (β = 0.12 ± 0.04, p = 0.003), and mean temperature (β = 0.35 ± 0.13, p = 0.006). In contrast, diurnal temperature range at short temporal lags (7-14 days; M185, M214; deviance explained: 41.7-42.4%) was negatively associated with population growth (β = –0.51 to –0.55, p = 0.002-0.011).

For the female *Ae. aegypti* population, precipitation at 21– and 28-day lags were identified as the best-fit single predictors in univariate models (M132-M133; AIC = 135.8-135.9; deviance explained: 43.7-44.0%), showing a consistent positive effect (β = 0.02 ± 0.01, p = 0.007-0.010). Relative humidity and mean temperature at similar temporal lags also showed positive associations in the best-supported bivariate models (M125; deviance explained: up to 51.2%). Conversely, diurnal temperature range at short lags (7-14 days; M135) was negatively associated with population growth. For the male population, precipitation, humidity, and temperature at 14 and 28-day lags were identified as predictors in the best-supported models (M164; deviance explained: up to 61.6%) (**S5 Fig, S4 File**).

For *Cx. quinquefasciatus*, models incorporating precipitation and temperature at 21 and 28-day lags (M9, M11; deviance explained: 32.7-34.8%) provided best-fit support (ΔAIC < 2), suggesting that mosquito abundance increases with precipitation but decline with temperature fluctuations. When assessing the male mosquito population alone, none of the evaluated climatic variables showed significant associations (ΔAIC < 2; all p > 0.1) whilst explanatory power remained low (pseudo-R^2^ < 14%). Multiple competing models showed similar AIC support, indicating uncertainty regarding the relative contribution of individual climatic variables (**S6 Fig**, **S4 File**), and limited effects on male mosquito abundance[79]. Overall, the association observed between climatic variables and local mosquito abundance support their utility as potential indicators of local vector population dynamics [32].

### A DENV-3 positive mosquito pool

No pathogenic flaviviruses known to be transmitted in Mexico by *Culex* species (*i.e.,* WNV and SLEV) [80,81] were detected using the initial pan-flavivirus PCR screening [82]. In contrast, a single pool of *Ae. aegypti* females (n=50) collected in October 2022 (epidemiological week 43) tested positive for DENV by PCR, with pool collection date coinciding with the peak in new dengue cases reported for both Yucatan State and Merida City (**Fig. 2a**) [83]. We estimated a Minimum Infection Rate (MIR) of 0.28 per 1,000, while the estimated Maximum Likelihood Estimate (MLE) was 0.29 per 1,000 (95% CI: 0.02-1.25). Our MIR/MLE estimates fall within the same order of magnitude as historic viro-entomological surveillance estimates from Mexico (∼1 per 1,000 female mosquitoes) [31]. Comparably, our estimates were marginally higher than the mean MIR value obtained through the TIRS trial during 2023 (MIR: 0.0765; CI 95%: 0.0585-0.0981) (*Puerta-Guardo et al. unpublished*) [31,40].

**Fig 2.**
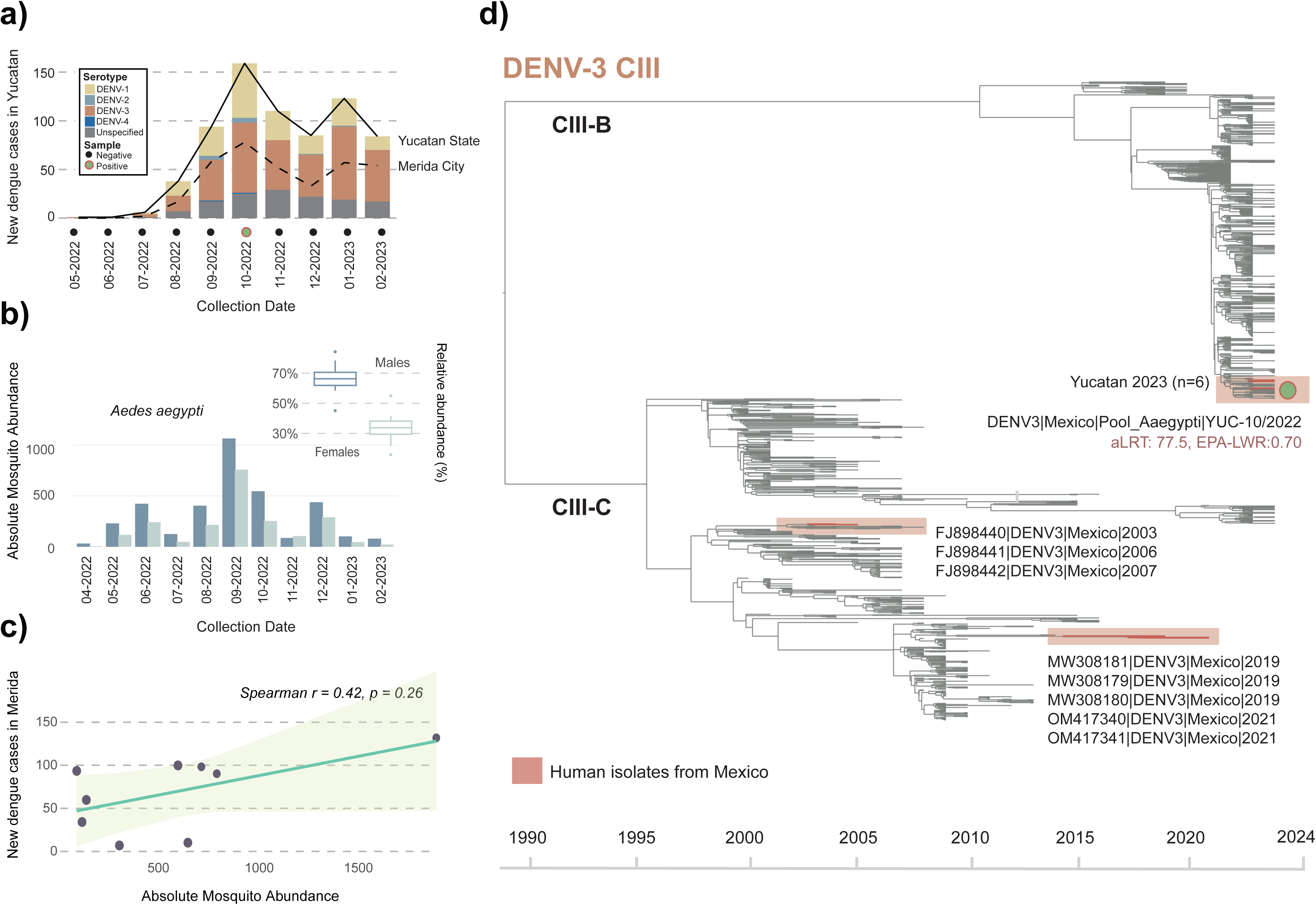
Detection of DENV-3 genotype III American II lineage from *Ae. aegypti* mosquitoes collected in the market. (a) Epidemiological trends of dengue in Yucatan State and Merida City recorded during May 2022 to February 2023 derived from official surveillance data. Bars represent new reported dengue cases by serotype (DENV-1,2,3 and 4). The timing of mosquito sampling within the market is indicated below, with a coloured circle denoting a single virus detection event (black = negative; green circle with orange outline = positive pool). (b) Monthly absolute abundance of *Ae. aegypti* mosquitoes within the market. Bars indicate total counts of male and female individuals. The boxplot shows the relative abundance by sex ratio across the full sampling period, with females consistently representing the majority. (c) Correlation between absolute *Ae. aegypti* abundance in the market and new dengue cases reported in Merida City. (d) Time-scaled ML tree showing the phylogenetic placement of the DENV3|Mexico|Pool_Aaegypti|Mex-Yuc|10/2022 partial sequence recovered from a mosquito pool sampled in October 2022 (indicated with a green circle with orange outline). DENV3|Mexico|Pool_Aaegypti|Mex-Yuc|10/2022 falls within the genotype III American II lineage (clade III-B), displaying moderate branch support (aLRT = 77.5, EPA-LWR = 0.70). Other human-derived isolates from Yucatan collected in 2023 are distantly related, whilst earlier human-derived isolates from elsewhere in Mexico collected between 2003-2021 fall within clade III-C.

DENV detection occurred during a period of known transmission, with the temporal overlap between the positive sample and the peak in dengue cases suggesting concordance between the local circulation of infected vectors at the market and the rise in dengue cases at the city level (**Fig. 2b**). A weak positive correlation was observed between reported dengue cases in Merida City and *Ae. aegypti* abundance sampled in the market. However, the association was not statistically significant (Spearman r = 0.23, p = 0.52). We further evaluated whether the association changed when comparing *Ae. aegypti* abundance from the previous month. Introducing a one-month lag increased the correlation strength (Spearman r = 0.42, p = 0.265; n = 9), although it remained non-significant (**Fig. 2c**). Although the small sample size limits statistical power, the pattern observed between absolute vector abundance (September 2022) and the subsequent peak in dengue cases (October 2022) is consistent with the expected lag in dengue transmission dynamics, where the expansion of infected vector populations typically precedes increases in reported cases [84]. Trap effort, temporal mosquito abundance, and climatic variables were further interpreted in the context of the outcomes derived from the TIRS trial [40] (for extended results, see **S2 Text and S2 Fig**).

### Detection of DENV-3 genotype III American II lineage (CIII-B) in Yucatan, Mexico

Our semi-nested PCR assay was successful to obtain partial NS5 (941 nt) and Env (296 nt) fragments from our DENV-positive mosquito pool (named here DENV3|Mexico|Pool_Aaegypti|Mex-Yuc|10/2022, **S5 File**). However, low viral RNA yield prevented full-genome sequencing, as has been noted in previous studies [85]. BLAST analysis of the concatenated and individual partial sequences revealed consistent results, with hits matching virus isolates assigned to the DENV3 3III_B.3.2 lineage from the Americas sampled during 2022 (as an illustrative example: DENV3/Cuba/2022-09-05/Yale-DN00829. Accession: OQ821537, Score: 1707 bits, E-value: 0.0, Identity: 99%, Coverage: 100%).

We further assessed the phylogenetic placement of the concatenated 1236 nt sequence amongst other DENV-3 genomes representing Clade III diversity circulating in North, Central and South America sampled between 2000 to 2024 (**S2 File**). Congruently, phylogenetic placement showed that DENV3|Mexico|Pool_Aaegypti|Mex-Yuc|10/2022 grouped with other viral genomes assigned to the 3III_B.3.2 lineage sampled from the USA and the Caribbean in 2022-2023 [36] (**Fig. 2d, S6 File**). Overall placement was consistent in both the full phylogenetic reconstruction and using EPA, with moderate branch support values (aLRT/LWR > 0.70). The six DENV-3 genomes from human cases derived from the TIRS trial [40] collected in early 2023 from Merida City also fall within 3III_B.3.2 lineage close to DENV3|Mexico|Pool_Aaegypti|Mex-Yuc|10/2022, but are not directly related. Other earlier DENV-3 genomes from Mexico publicly released by 2022 fall within subclade CIII-C. Though this hypothesis would require formal testing entailing expanded datasets under a phylogeographic framework [19], our phylogenetic analysis suggests a likely introduction into Yucatan through the Caribbean corridor.

### Wastewater surveillance: detection of SARS-CoV-2

We further assessed the temporal association between SARS-CoV-2 RNA detection from wastewater at the market and COVID-19 cases reported in Merida City and broader Yucatan state. During the study period, two epidemic peaks were officially recorded by the National Ministry of Health [86]: June-July 2022 (epidemiological weeks 25-28, representing the largest transmission peak), and November-December 2022 (epidemiological weeks 48-51). Sampling spanned both infection waves, during which viral RNA was detected across both peaks, suggesting a mirrored COVID-19 case incidence in the market (**Fig. 3a**). Overall, Ct-values were higher (>35) during the first infection wave, indicating lower viral RNA concentration in wastewater despite higher transmission. However, this observation is confounded by a lower sampling intensity during the earlier period. Conversely, lower Ct-values (<35) recorded during the second infection wave indicate a closer temporal alignment with new case reports. Samples collected during epidemiological weeks with few or no new cases displayed higher Ct-values (>37), or no amplification at all (**Fig. 3b**).

**Fig 3.**
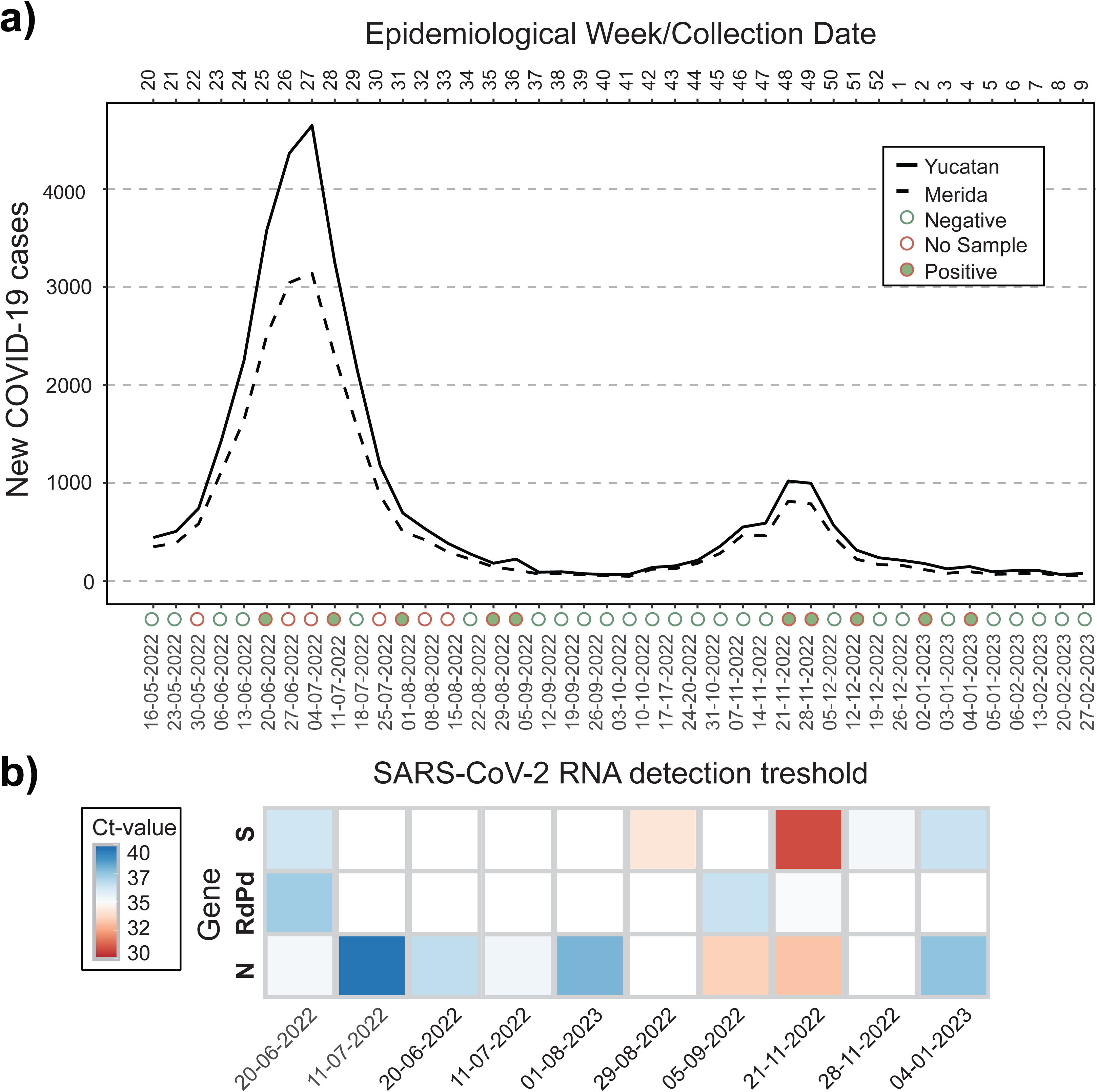
Detection of SARS-CoV-2 RNA from wastewater in association with new COVID-19 cases in Yucatan. (a) Weekly new COVID-19 cases in Yucatan State (solid line) and Merida City (dashed line) reported between May 2022 and February 2023, reflecting the official epidemiological calendar. Wastewater collection overlapped both epidemic peaks recorded during the sampling period. Circles indicate wastewater sampling status. Green outlined circles show negative samples, red outlined circles indicate no sample taken, and the green circles with orange outline correspond to positive samples. (b) Heatmap for SARS-CoV-2 RNA detection from wastewater samples using qPCR, showing cycle threshold (Ct) amplification values for the N, RdRp, and S virus genes across collection dates. Lower Ct-values (30-33) coincide with an increase in COVID-19 cases during the second epidemic wave. In contrast, higher Ct-values (>37) or no amplification were observed during low case incidence periods.

We further modelled the probability of detecting SARS-CoV-2 RNA as a function of weekly COVID-19 case count. Higher weekly case counts were associated with an increased probability of wastewater detection (OR = 1.36 per 500 cases; 95% CI: 0.93-1.99; p = 0.095), corresponding to around 36% increase in odds of detection per 500 additional reported cases. Model comparison identified a two-week lag (Lag 2) as the best-fitting model based on AIC (AIC = 40.14). Correlation analyses similarly showed an increase from Lag 0 (ρ = 0.36, p = 0.029) to Lag 2 (ρ = 0.49, p = 0.003) (**S4 Table, S7 Fig**). However, the association was not statistically significant, indicating limited power due to sparse sampling. Thus, our results do not provide evidence for a lead time of wastewater detection relative to reported cases during high-transmission periods.

### Live bird surveillance: detection of AIVs

A total of 21 cloacal swabs from birds sold at the Lucas de Gálvez market were screened using RT-qPCR targeting the influenza A virus matrix (M) gene. Two juvenile turkeys (*Meleagris gallopavo*) from the same cage tested positive with high Ct values (38.96-39.43). Subtype-specific assays for H5 and H7 were negative, suggesting either infection with a non-H5/H7 avian influenza A virus or insufficient viral RNA for subtyping. Subsequent BLAST analyses yielded best hits to AIVs (*e.g.,* strain A/H5N2/chicken/Mexico/28159-541/1995; Accession: GU052693; Score: 64 bits; E-value: 4e-08; Coverage: 10%). However, a low sequence coverage limited taxonomic assignment, and findings should be interpreted cautiously. Nevertheless, these results are consistent with a possible cryptic circulation of AIVs in birds within market settings, and are in line with the known ecology of avian influenza viruses and recent reports of avian influenza infections in Yucatan and broader Mexico [25]. Thus, they further support the relevance of a continued monitoring of AIVs in domestic birds following a One Health framework [87].

## DISCUSSION

We implemented a virus monitoring strategy within a traditional urban market in Merida City, Yucatan, Mexico. Our key findings include an early identification of DENV-3 lineage III_B.3.2 in a pool of *Ae. aegypti* mosquitos, SARS-CoV-2 RNA detection from wastewater mirroring broad epidemiological trends recorded at a municipal level, and putative signals of a cryptic AIV circulation in live birds sold at the market.

Within the entomological component of our study, the joint analysis of mosquito population structure with climatic variables provided an ecological context for arbovirus circulation at a local scale. In Mexico, environmental and entomological indicators are incorporated into epidemiological surveillance to estimate the risk of transmission for arboviruses [31,33,65]. However, while these provide an important contextualisation, they remain indirect proxies of virus circulation and transmission. As an example, evidence from the TIRS trial showed that a substantial decrease in *Ae. aegypti* household density did not correspond to a reduction in symptomatic arboviral disease incidence [40], highlighting the disconnect between vector and virus dynamics. Thus, integrating signals from molecular data directly derived from virus monitoring can contribute to a better understanding of virus circulation in vector populations.

Although based on a single positive mosquito pool, our result further highlights how molecular data can capture viral genetic diversity not recovered through classic epidemiological surveillance. Beyond standard virus serotyping, full genome sequences or fragments can support the characterisation of virus population dynamics, as we show for the recent introduction and spread of DENV-3 lineage III_B.3.2 in Mexico [39]. However, similar to other local viro-entomological monitoring efforts, achieving high virus detection rates would require resource-intensive mosquito sampling and screening strategies. Furthermore, as reflected by some aspects of our experimental design, a broader spatial and temporal framework would be necessary for successful long-term implementation. Thus, future studies should incorporate larger numbers of sampling sites and traps, simultaneous and standardised surveillance across multiple settings (*e.g.,* markets and households), longer temporal series, and spatial-temporal analyses to assess synchrony and predictive capacity.

The wastewater component of our study also illustrates potential limitations of environmental sampling for capturing early signals of viral transmission, as we found no clear evidence of a lead time relative to reported cases during peak transmission periods. Nonetheless, the temporal overlap noted between the detection of SARS-CoV-2 RNA and the COVID-19 epidemic waves reported supports the use of wastewater for monitoring ongoing viral transmission, comparable to other studies using similar strategies implemented at different scales [71]. However, virus detection sensitivity from wastewater surveillance critically depends on a systematic and temporally-resolved sampling, where municipal facilities may be better suited to capture virus transmission dynamics over time.

Sampling constraints observed within all monitoring components of our pilot resulted in important biases that significantly reduced statistical power. Importantly, these also reflect logistical complexities that underscore the need for sustained collaboration with local authorities and the communities involved for these surveillance strategies [88]. Importantly, the interpretation of temporal fluctuations in viral circulation should also consider the social and behavioural factors that influence human activity within sampling sites. Although such data were not available for the market, integrating human mobility patterns, together with anonymised reporting of febrile illness followed by serological and molecular diagnostics. Altogether, these could provide high-resolution insight into local virus transmission associated with social dynamics and human behaviour, improving our current understanding of the broader drivers of transmission.

Despite limitations, our results suggest that even under sparse sampling, monitoring viruses of epidemiological relevance at this urban site was feasible, highlighting an added value of integrating molecular data within classic epidemiology surveillance, specially where genomic capacity remains limited [89]. While urban markets represent only one of several possible scenarios for similar implementations, our approach provides a focused perspective on a particular site where viral circulation coincided with high human activity and potential human-animal interactions. A prompt detection of viruses of epidemiological relevance remains important to enable proactive control strategies that can help reduce public health burden and economic loss, particularly important for mosquito-borne diseases [89]. With improved sampling consistency and expanded sequencing capacity, similar approaches could contribute to strengthening existing surveillance efforts and inform context-specific public health responses. At an estimated low operational cost, this strategy represents an economically feasible complementary effort [90].

## DATA AVAILABILITY

The partial DENV-3 sequence generated in this study is available in **S5 File**. The corresponding phylogenetic ML tree is available in **S6 File**.

## ETHICAL CONSIDERATIONS

All animal handling protocols were approved by the Institutional Bioethics Subcommittee for the Care and Use of Experimental Animals, UNAM (SICUAE.DC-2022/2-4). Experimental design for this study was approved under the Bioethics Committee of the Universidad Autonoma de Yucatan, UADY (CB-CCBA-I-2021-002).

## ACKNOWLEDGMENTS

This study was supported by the John Fell OUP Research Grant ATD00390 (M.E.Z and M.U.G.K), the Wellcome Infectious Disease Award (317324/Z/24/Z to M.G.K, H.P.G and M.E.Z), the Secretaria de Ciencia, Humanidades, Tecnología e Inovación award (SECIHTI, Mexico) through the PRONACES Health grant (PRONAII project number 303002, G.S) and the Ciencia Básica y de Frontera programme (CBF2023-2024-3184, M.G.K), and the UKRI Innovation BSRC/EPSRC/NIHR 971557 grant (A.R.S). M.G.K is funded through a Sanger International Fellowship award. M.E.Z is funded by a UCL Rosetrees Excellence Fellowship UCL2024\2. P.M.D was funded through the doctoral program at ‘Posgrado en Ciencias de la Produccion y de la Salud Animal-UNAM’ through the SECIHTI doctoral scholarship. M.U.G.K. acknowledges funding from The Rockefeller Foundation (PC-2022-POP-005), Health AI Programme from Google.org, the Oxford Martin School Programmes in Pandemic Genomics & Digital Pandemic Preparedness, European Union’s Horizon Europe programme projects MOOD (#874850) and E4Warning (#101086640), Wellcome Trust grants 303666/Z/23/Z, 226052/Z/22/Z & 228186/Z/23/Z, the United Kingdom Research and Innovation (#APP8583), the Medical Research Foundation (MRF-RG-ICCH-2022-100069), UK International Development (301542-403), the Bill & Melinda Gates Foundation (INV-063472) and Novo Nordisk Foundation (NNF24OC0094346). B.G is further funded by Wellcome Trust grants 303666/Z/23/Z, 226052/Z/22/Z & 228186/Z/23/Z. The contents of this publication are the sole responsibility of the authors and do not necessarily reflect the views of the European Commission or the other funders. The funders had no role in study design, data collection and analysis, decision to publish, or preparation of the manuscript.

We would like to acknowledge the community and staff of the Lucas de Galvez and San Benito markets, particularly to Freddy Francisco Arjona Candila, Carlos Manuel Euan Canul, and Santos José Noh Sierra for their support in to sample logistics and collection. We thank Roger Arana and Laura I. López Apodaca for their support in sample collection. We thank Julia Leleu, Ivonne Mora Herrera, and Cesar Cañas-Alamilla for their technical assistance in the laboratory. Data from the TIRS trial and DENV-3 sequences from Yucatan 2023 were provided by Dr. Puerta-Guardo and Azael Che-Mendoza (UADY).

