## Supplementary Information for "“A Market-Based Virus Monitoring Strategy Complementary to Epidemiological Surveillance in Urban Settings”"

**CONTENT**

**Supplementary Text**

- **S1 Text.** Contextual epidemiology and disease dynamics of DENV in Mexico and Latin America
- **S1 Fig.** Dengue burden and genomic surveillance across Latin America
- **S2 Text.** Exploratory comparison of mosquito abundance dynamics between market and household surveillance from TIRS
- **S2 Fig.** Comparison of temporal mosquito abundance dynamics between market and household surveillance

**Supplementary Tables**

- **S1 Table.** Mosquito sampling effort and population structure
- **S2 Table.** Variance inflation factor (VIF) for climatic data
- **S3 Table.** Logistic regression for COVID-19 cases and SARS-CoV-2 RNA detection in wastewater

**Supplementary Figures**

- **S3 Fig.** Live bird sampling scheme
- **S4 Fig.** Relative mosquito abundances and sex ratios
- **S5 Fig.** GLMs for *Ae. aegypti*
- **S6 Fig.** GLMs for *Cx. quinquefasciatus*
- **S7 Fig.** Predicted probability of SARS-CoV-2 RNA detection in wastewater

**Supplementary Files**

- **S1 File.htlm** Interactive live bird sampling layout
- **S2 File.xlsx** Metadata for DENV3 genomes used in phylogenetic inference
- **S3 File.microreact** Microreact Interactive visualization of the DENV-3 tree
- **S4 File.xlsx** GLM outputs
- **S5 File.fasta** DENV3|Mexico|Pool_Aaegypti|Mex-Yuc|10/2022
- **S6 File 6.tree** DENV3 ML phylogenetic tree

**S1 Text. Contextual epidemiology and disease dynamics of DENV in Mexico and Latin America**

Dengue surveillance data in Latin America shows that although case reporting and laboratory-confirmed diagnosis capacity are substantial, genomic surveillance remains comparatively limited and heterogeneous across countries. Specifically, while dengue case reporting strongly correlates with PCR-confirmed cases (R = 0.88), the relationship between case counts and publicly available viral genomes is weak (R = 0.31), indicating an overall gap in virus sequencing. Within this landscape, Mexico operates an highly functional epidemiological surveillance system integrating data from over 16,000 health facilities nationwide, making it one of the strongest systems in Latin America. However, when it comes to sequencing capacity, Mexico occupies an intermediate position (**Figures 1 and 2**). Nonetheless, with an increasing genomic sampling relative to disease burden [1,2], retrospective efforts have helped characterised historic DENV spatiotemporal dynamics over decades [3] and identified the recent spread of epidemiologically relevant lineages [4].

Since 2018, DENV-1 and DENV-2 serotypes dominated in Mexico, with a marked shift to DENV-1 dominance observed after 2019 [3]. Although DENV-3 was detected at low incidence levels from 2009 onwards, it only began rising in incidence after mid-2022, with full dominance observed after 2023 [5,6]. By 2025, DENV-3 national cases accounted for over 90% hospitalizations [7]. Accordingly, DENV-3 lineage 3III_B.3.2 was circulating in neighbouring regions and was later reported in Mexico, in association with the serotype shift recorded at a regional scale in 2022-2023 [4,8,9]. Serotype replacement is common in dengue epidemiology; however, serotype turnover does not imply an increase in epidemic intensity or clinical severity. The latter are likely driven by both viral genetics and population immunity structure, including seroreactivity and antibody-dependent enhancement dynamics [10,11]. In this light, it has been suggested that the most recent epidemic peaks observed in Brazil cannot be solely explained by a DENV-3 serotype replacement and expansion of lineage III_B.3.2 across the country [8].

Although further genomic and epidemiological data is needed to clarify the underlying drivers of the most recent epidemic shift observed in Mexico, this phenomenon was likely enabled by both changes in the immune structure of the population, coupled with a virus lineage replacement event. Supporting this observation, our phylogenetic analysis shows that DENV-3 isolates from Mexico sampled between 2019 and 2021 belong to clade III-C, indicating prior circulation of this endemic lineage to which a fraction of the population likely have had some immunity. However, a recent serological survey amongst children aged 12-15 in Merida (data derived from the TIRS trial [12] revealed that DENV-3 seropositivity was minimal prior to 2023, suggesting limited or no immunity within this group (*Puerta-Guardo et al. unpublished*). Therefore, the subsequent introduction of lineage III_B.3.2 could have facilitated epidemic expansion of DENV-3 due to a limited prior exposure of vulnerable populations [9].

Our analysis suggests that the introduction of lineage III_B.3.2 into Yucatan might have occurred through the Caribbean corridor as early as October 2022. This observation is consistent with a pairwise similarity/close phylogenetic placement of our mosquito-derived sequence and other III_B.3.2 viruses circulating in the Caribbean and North America collected during 2022 [13], all of which precede cases from Yucatan and wider Mexico [4]. Nonetheless, testing this hypothesis would entail a detailed phylogeographic analysis integrating newly generated data from Mexico and neighboring geographic regions, ideally incorporating both mosquito- and human-derived virus genomes, as they become available [3].

### S1 Fig. Dengue burden and genomic surveillance across Latin America

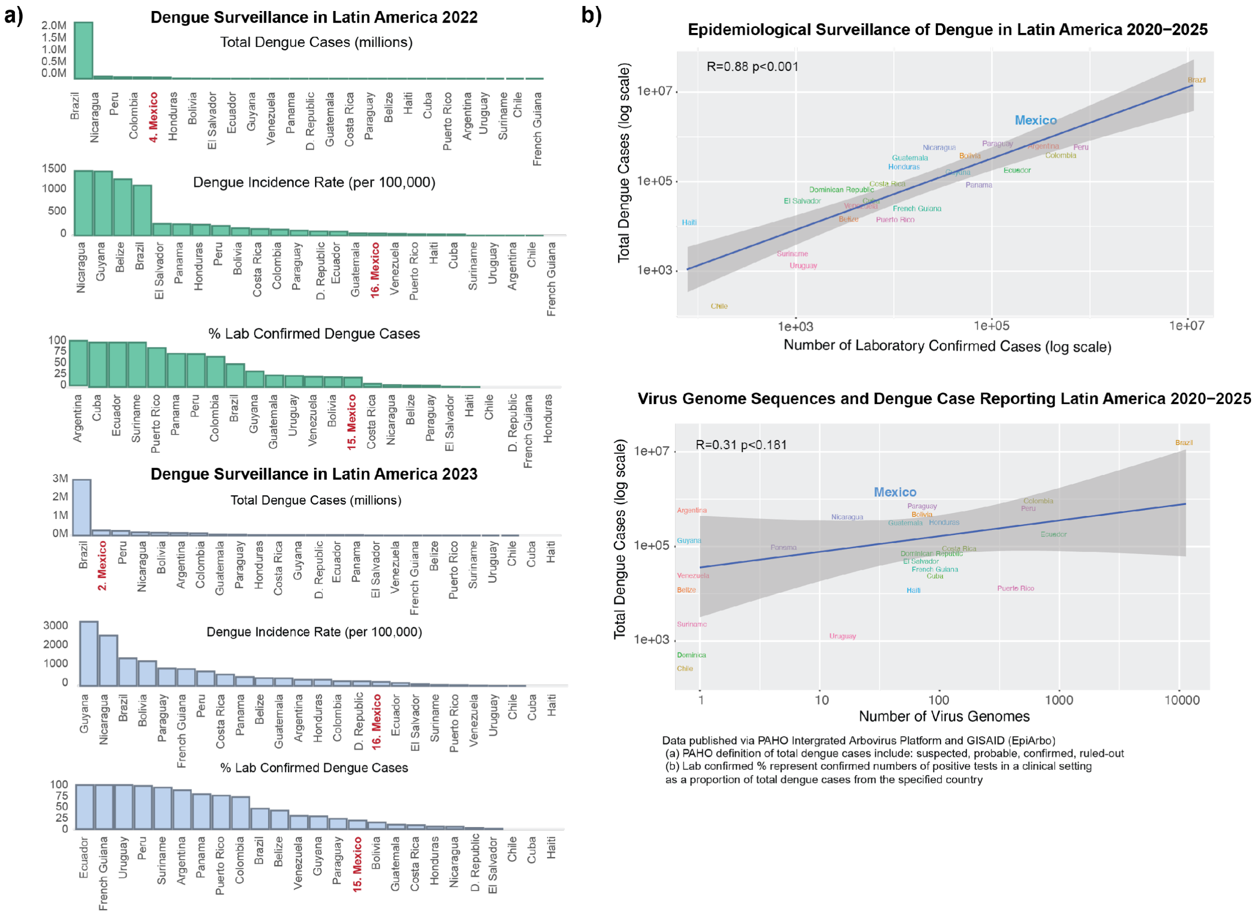


Data published via PAHO Intergrated Arbovirus Platform.
(a) PAHO definition of total dengue cases include: suspected, probable, confirmed, ruled-out
(b) Incidence rates are cumulative of all dengue cases per 100,000 population, as of latest epi week of 2022.
(c) Lab confirmed % represent confirmed numbers of positive tests in a clinical setting as a proportion of total dengue cases from the specified country

Comparison of dengue burden, laboratory confirmation, and genomic surveillance across countries in Latin America and the Caribbean. a) Left panels show country-level rankings for (i) total dengue cases, (ii) dengue incidence rate per 100,000 population, and (iii) percentage of laboratory-confirmed cases, presented for two representative periods (2022 and 2023). Mexico is highlighted in red to indicate its relative position across metrics. b) Right panels show correlations between (top) total dengue cases and number of laboratory-confirmed cases, and (bottom) total dengue cases and number of available viral genomes from GISAID

**S2 Text. Exploratory comparison of mosquito abundance dynamics between market and household surveillance from TIRS**

To further contextualise the market-based entomological observations, exploratory analyses were conducted by comparing temporal fluctuations in *Ae. aegypti* abundance recorded at the market with those observed in household surveillance during the TIRS trial [12]. Standardised monthly abundance patterns were visualised using temporal trend plots, further applying Spearman correlation (to evaluate the predictive association between both datasets.

Although some temporal overlap was observed during periods of increased mosquito abundance, patterns were inconsistent across the sampling period with no statistical significance (Spearman $\rho = 0.5$, p-value > 0.1) (**S2 Fig**). Thus, we did not identify evidence for synchrony, a temporal lag structure, or a predictive association between mosquito abundance observed within the market and that recorded in residential settings.

However, due to substantial differences in sampling design and statistical power across datasets, these findings should be interpreted cautiously. Household surveillance conducted through TIRS included systematic mosquito collections through individual aspirations across approximately 750 households per month [12], whereas the market-based mosquito surveillance relied on trap use, with fewer trapping points over a shorter temporal series, which limits such comparison.

Our results indicate that the current dataset is insufficient to determine whether market-based mosquito surveillance can reliably capture or predict broader city-scale vector dynamics. Moreover, this market represents a localised ecological microenvironment that may not necessarily reflect broader residential transmission dynamics occurring across the city. Future studies incorporating larger longitudinal datasets, standardised parallel sampling designs, and expanded spatial coverage will be required to formally evaluate this.

**S2 Fig. Comparison of temporal mosquito abundance dynamics between market and household surveillance**

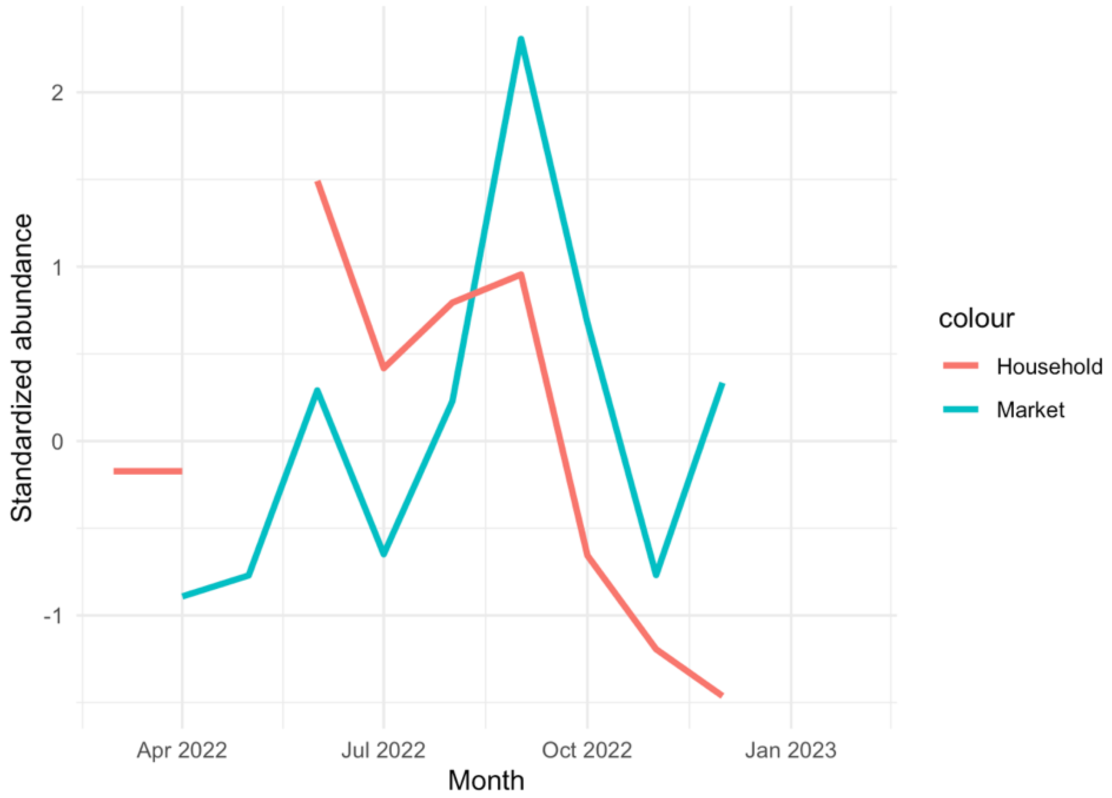


Standardised monthly abundance patterns of *Ae. aegypti* recorded from market-based surveillance and household collections conducted during the TIRS trial in Merida City between 2022-2023. Standardised abundance facilitates a temporal comparison between surveillance systems with different sampling intensities. Both datasets show partial temporal overlap during periods of increased mosquito abundance, although no consistent synchrony or a predictive association was supported.

**S1 Table.** Mosquito sampling effort and population structure

| Year | Month | Sampling Period Epidemiological week | Trap hours | Total Individuals Collected | *Culex quinquefasciatus* **(n=14,357/19,998; 71.8%)** | | | | | *Aedes aegypti* **(n=5,641/19,998; 28.2%)** | | | | |
|---|---|---|---|---|---|---|---|---|---|---|---|---|---|---|
| | | | | | **Female*** | | | **Male** | | **Female*** | | | **Male** | |
| | | | | | **f/g** | **e** | **%** | | **%** | **f/g** | **e** | **%** | | **%** |
| 2022 | Apr | 17 | 240 | 68 | 2 | 16 | 26.47 | 12 | 17.64 | 11 | 21 | 8.82 | 6 | 47.05 |
| | May | 18 | 840 | 668 | 13 | 130 | 21.40 | 178 | 26.64 | 40 | 190 | 17.51 | 117 | 34.43 |
| | | 19 | | | | | | | | | | | | |
| | | 20 | | | | | | | | | | | | |
| | Jun | 26 | 360 | 1,432 | 0 | 407 | 28.42 | 359 | 25.06 | 5 | 419 | 16.89 | 242 | 29.60 |
| | Jul | 30 | 360 | 1,113 | 7 | 520 | 47.34 | 413 | 37.10 | 2 | 123 | 4.31 | 48 | 11.23 |
| | Aug | 34 | 360 | 1,523 | 8 | 532 | 35.45 | 363 | 23.83 | 8 | 396 | 14.18 | 216 | 26.52 |
| | Sep | 38 | 360 | 5,857 | 1 | 2847 | 48.62 | 1185 | 20.23 | 10 | 1055 | 12.95 | 759 | 18.18 |
| | Oct | 43 | 360 | 2,224 | 3 | 955 | 43.07 | 465 | 20.90 | 1 | 546 | 11.42 | 254 | 24.59 |
| | Nov | 48 | 360 | 694 | 17 | 318 | 48.27 | 166 | 23.91 | 0 | 87 | 15.27 | 106 | 12.53 |
| | Dec | 50 | 360 | 2,165 | 2 | 941 | 43.25 | 493 | 22.77 | 5 | 433 | 13.44 | 291 | 20.23 |
| 2023 | Jan | 3 | 360 | 408 | 8 | 183 | 46.81 | 69 | 16.91 | 4 | 98 | 11.27 | 46 | 25 |
| | Feb | 9 | 360 | 3,846 | 4 | 2237 | 58.26 | 1503 | 39.07 | 0 | 80 | 0.57 | 22 | 2.08 |
| | | **TOTAL** | **4,320** | **19,998** | **65** | **9,086** | **63.7%** | **5,206** | **36.3%** | **86** | **3,448** | **62.7%** | **2,107** | **37.4%** |

*Female mosquitoes were pooled by feeding status (f = fed, g = gravid, e = empty), with relative percentages indicating the proportion over the total sample per sampling period.

#### S3 Fig. Live bird sampling scheme

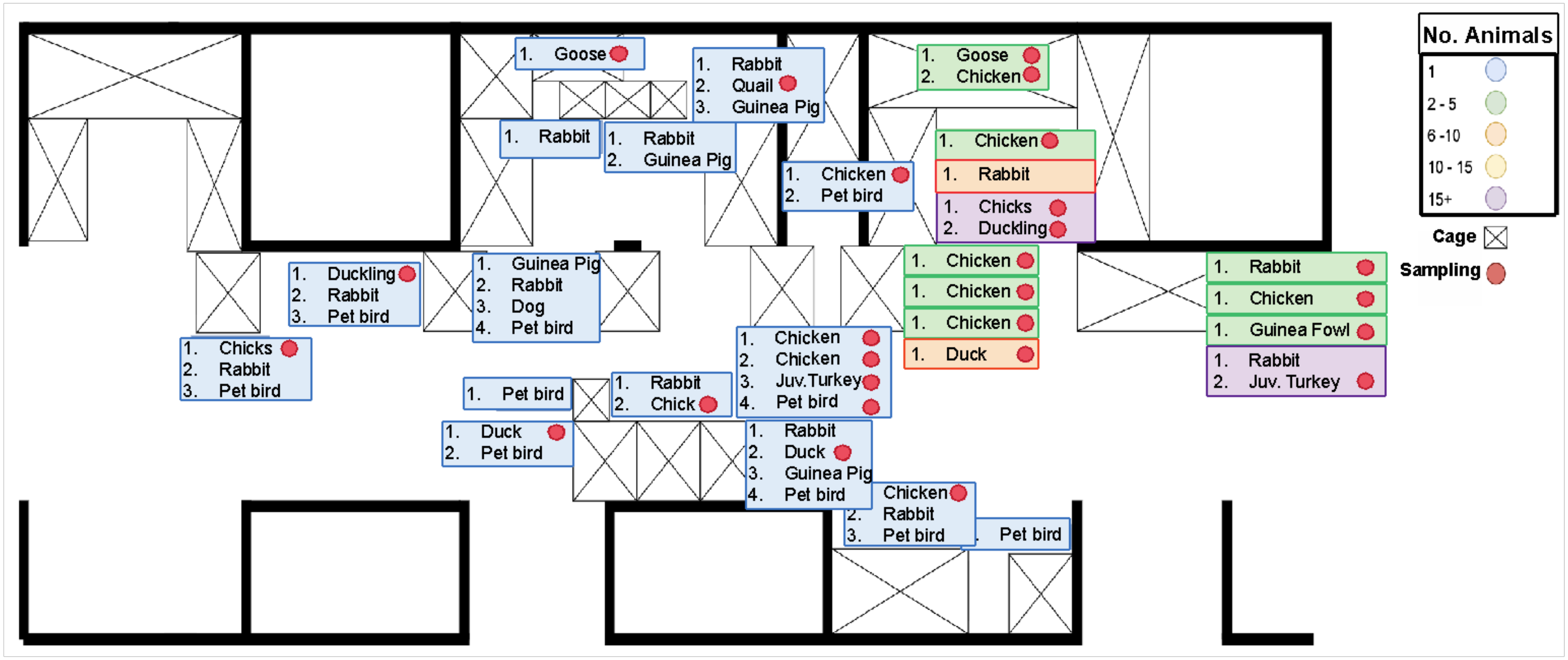


Schematic representation of the live animal vending facilities at the Lucas de Galvez Market, where a single sampling event of live bird took place. Cloacal swabs were collected from 21/70 (30%) of all birds within accessible cages. Sampled species included juvenile turkeys (*Meleagris gallopavo*), adult and juvenile chickens (*Gallus gallus domesticus*), adult and juvenile ducks (*Anas platyrhynchos domesticus*), one guinea fowl (*Numida meleagris*), and a goose (*Anser anser domesticus*). Colored rectangles indicate the number of animals per cage (marked in an X, with red circles indicating cages from which samples were taken. An interactive visualization of the sampling layout is provided in **Supplementary File 1**.

**S4 Fig.** Relative mosquito abundances and sex ratios

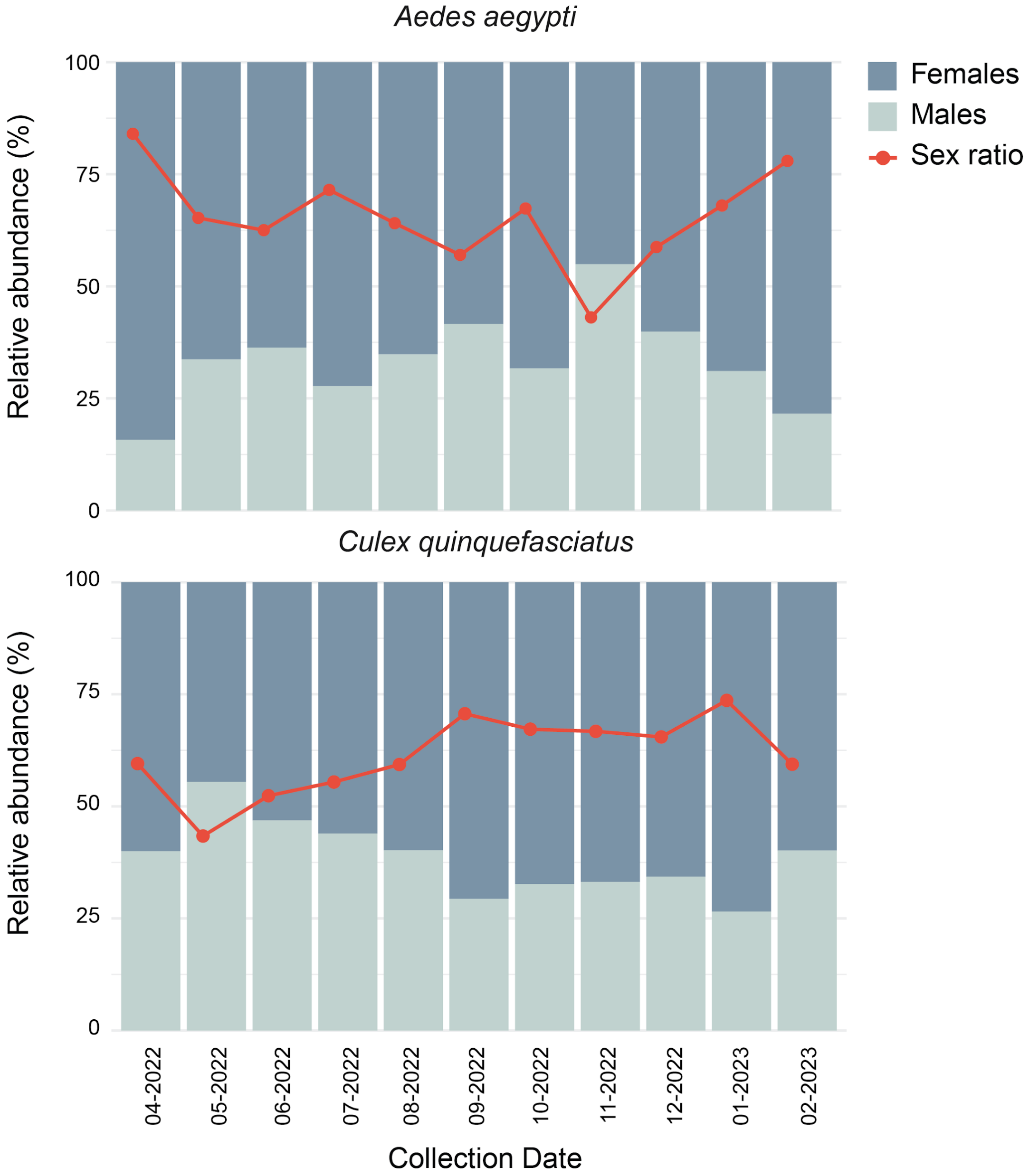


Stacked bar plots show the relative monthly abundance (%) of female (dark blue) and male (light blue) mosquitoes collected between April 2022 and February 2023. The red line indicates the sex ratio (female to male proportion) observed during each sampling period. *Ae. aegypti* (top) displayed greater variability in sex ratio over time, with female-biased peaks during April, September, and January. *Cx. quinquefasciatus* (bottom) showed a more stable sex ratio across months, with moderate female predominance.

**S1 Table.** Correlation matrix with variance inflation factor (VIF) for climatic data

| Variables * | GMT_7d_°C | RH_7d_% | Ppt_7d_mm | Variable | VIF |
|---|---|---|---|---|---|
| **GMT_7d_°C** | 1 | 0.43 | 0.32 | GMT_7d_°C | 1.272 |
| **RH_7d_%** | 0.43 | 1 | **0.92** | RH_7d_% | 7.662 |
| **Ppt_7d_mm** | 0.32 | **0.92** | 1 | Ppt_7d_mm | 7.005 |
| | **GMT_14d_°C** | **RH_14d_%** | **Ppt_14d_mm** | **Variable** | **VIF** |
| **GMT_14d_°C** | 1 | 0.06 | 0.22 | GMT_14d_°C | 1.130 |
| **RH_14d_%** | 0.06 | 1 | **0.86** | RH_14d_% | 4.061 |
| **Ppt_14d_mm** | 0.22 | **0.86** | 1 | Ppt_14d_mm | 4.254 |
| | **GMT_21d_°C** | **RH_21d_%** | **Ppt_21d_mm** | **Variable** | **VIF** |
| **GMT_21d_°C** | 1 | -0.1 | 0.34 | GMT_21d_°C | 2.665 |
| **RH_21d_%** | -0.1 | 1 | **0.84** | RH_21d_% | 7.943 |
| **Ppt_21d_mm** | 0.34 | **0.84** | 1 | Ppt_21d_mm | 8.881 |
| | **GMT_28d_°C** | **RH_28d_%** | **Ppt_28d_mm** | **Variable** | **VIF** |
| **GMT_28d_°C** | 1 | -0.2 | 0.29 | GMT_28d_°C | 2.115 |
| **RH_28d_%** | -0.2 | 1 | **0.77** | RH_28d_% | 4.850 |
| **Ppt_28d_mm** | 0.29 | **0.77** | 1 | Ppt_28d_mm | 5.086 |

* Climatic variables included geometric mean temperature (GMT, °C), mean minimum temperature (Tmin, °C), mean maximum temperature (Tmax, °C), mean diurnal temperature range (DTR, °C), cumulative relative humidity (RH, %), and cumulative precipitation (Ppt, mm)

**S5 Fig.** GLMs for *Ae. aegypti*

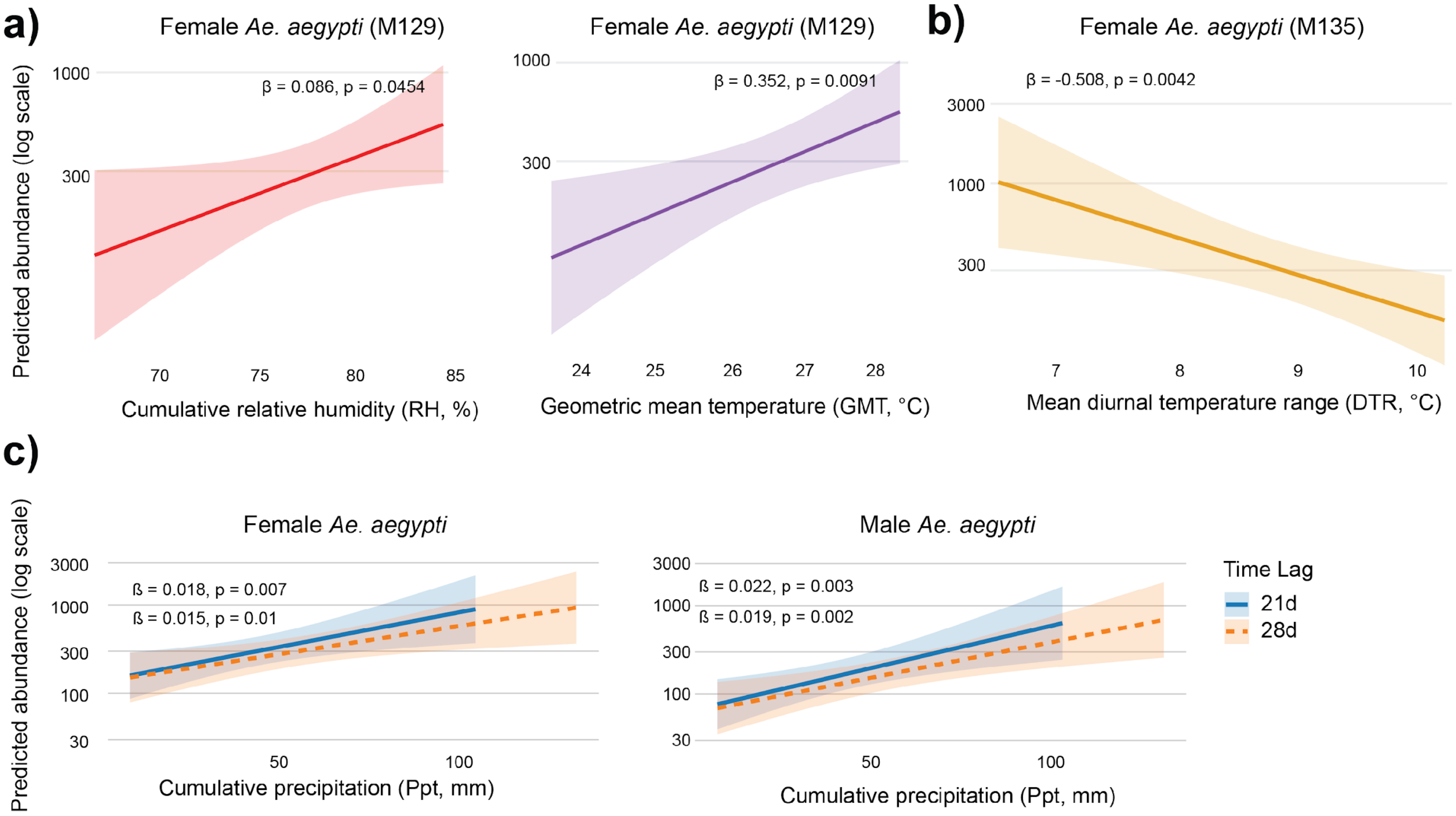


(a) Positive associations between cumulative relative humidity (RH, %) and geometric mean temperature (GMT, °C) and predicted female *Ae. aegypti* abundance (model M129). (b) Negative association between mean diurnal temperature range (DTR, °C) and female abundance (model M135). (c) Positive associations between cumulative precipitation (Ppt, mm) and predicted female (left) and male (right) *Ae. aegypti* abundance at 21-day (solid blue) and 28-day (dashed orange) lags. Shaded areas indicate 95% confidence intervals. Regression coefficients (β) and p-values are shown for each model. Predicted abundances are presented on a log scale.

**S6 Fig.** GLMs for *Cx. quinquefasciatus*

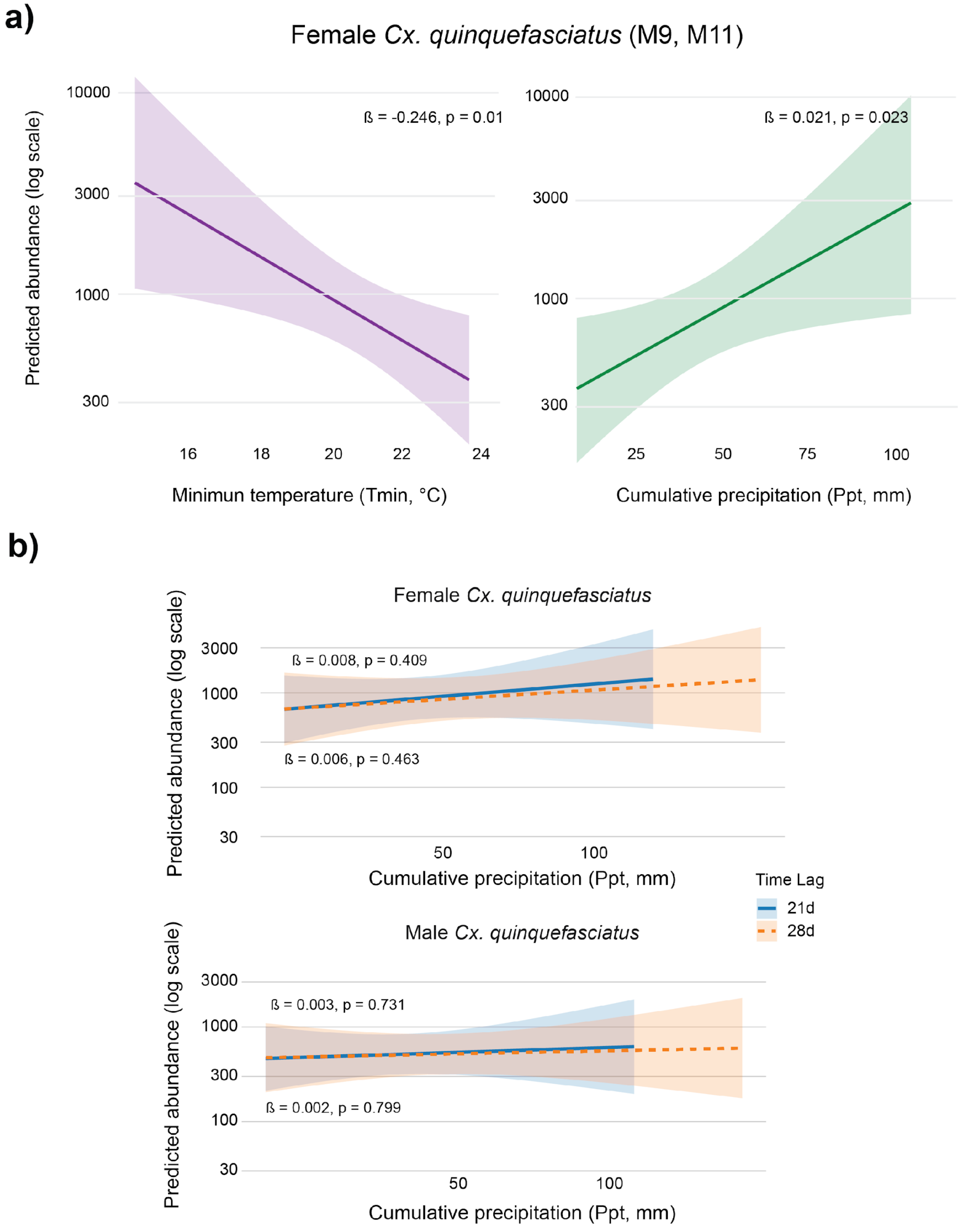


(a) Predicted female *Cx. quinquefasciatus* abundance showing a negative association with minimum temperature (Tmin, °C) and a positive association with cumulative precipitation (Ppt, mm) over a 21 days lag (Models M9, M11). (b) Predicted female (top) and male (bottom) *Cx. quinquefasciatus* abundance as a function of cumulative precipitation at 21-day (solid blue) and 28-day (dashed orange) lags. Shaded areas represent 95% confidence intervals. Regression coefficients (β) and *p*-values are shown for each model. Predicted abundances are presented on a log scale.

**S3 Table.** Logistic regression for COVID-19 cases and SARS-CoV-2 RNA detection in wastewater

| Lag (weeks) | OR per 500 new cases | 95% CI | p-value | AIC | Spearman ρ | p (Spearman) |
|---|---|---|---|---|---|---|
| Lag 0 | 1.41 | 0.96–2.07 | 0.066 | 41.47 | 0.364 | 0.029 |
| Lag 1 | 1.38 | 0.94–2.03 | 0.082 | 40.82 | 0.404 | 0.016 |
| Lag 2 | 1.36 | 0.93–1.99 | 0.095 | 40.14 | 0.494 | 0.003 |

**S7 Fig.** Predicted probability of SARS-CoV-2 RNA detection in wastewater

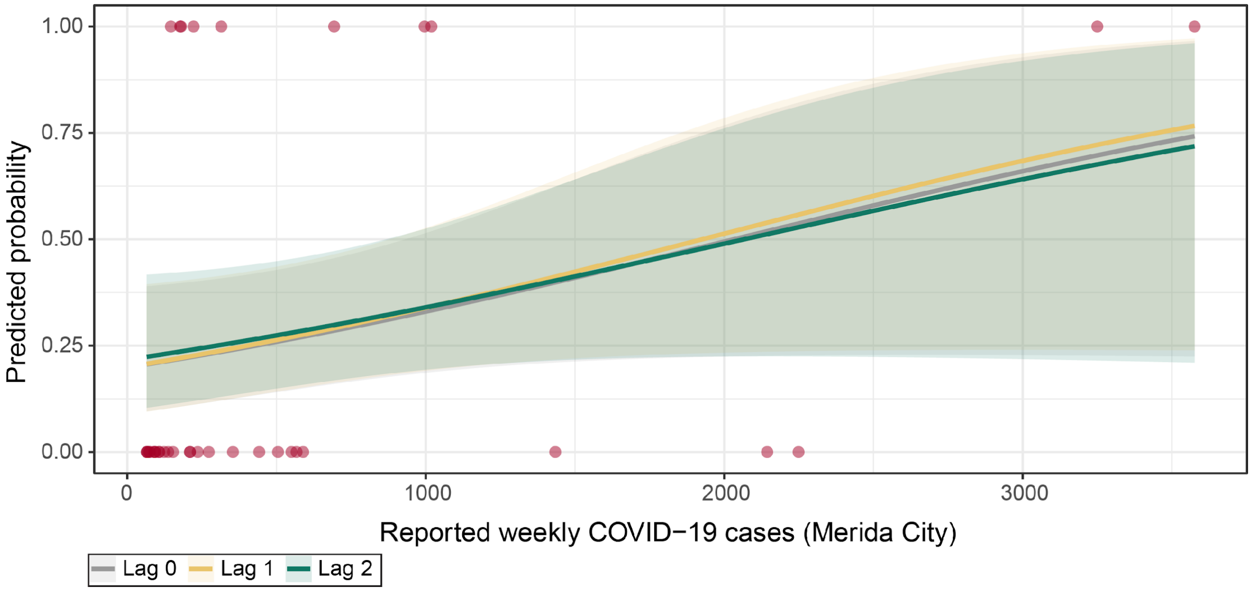


Predicted probabilities of SARS-CoV-2 RNA detection in wastewater as a function of weekly COVID-19 case counts in Merida city, estimated using Firth-penalized logistic regression models with 0–2-week temporal lags. Shaded areas represent 95% confidence intervals. Red points indicate observed wastewater detections (1 = positive, 0 = negative). Detection probability increased with rising case counts, with the two-week lag model (green) providing the best fit (AIC = 40.14), consistent with wastewater signals preceding reported cases.
